# Cov^2^MS: an automated matrix-independent assay for mass spectrometric detection and measurement of SARS-CoV-2 nucleocapsid protein in infectious patients

**DOI:** 10.1101/2022.02.09.22270547

**Authors:** Bart Van Puyvelde, Katleen Van Uytfanghe, Laurence Van Oudenhove, Ralf Gabriels, Tessa Van Royen, Arne Matthys, Morteza Razavi, Richard Yip, Terry Pearson, Marijn van Hulle, Jan Claereboudt, Kevin Wyndham, Don Jones, Xavier Saelens, Geert A. Martens, Christophe Stove, Dieter Deforce, Lennart Martens, Johannes P.C. Vissers, N. Leigh Anderson, Maarten Dhaenens

## Abstract

**INTRODUCTION:** The pandemic readiness toolbox needs to be extended, providing diagnostic tools that target different biomolecules, using orthogonal experimental setups and fit-for-purpose specification of detection. Here we build on a previous Cov-MS effort that used liquid chromatography-mass spectrometry (LC-MS) and describe a method that allows accurate, high throughput measurement of SARS-CoV-2 nucleocapsid (N) protein.

**MATERIALS and METHODS:** We used Stable Isotope Standards and Capture by Anti-Peptide Antibodies (SISCAPA) technology to enrich and quantify proteotypic peptides of the N protein from trypsin-digested samples from COVID-19 patients.

**RESULTS:** The Cov^2^MS assay was shown to be compatible with a variety of sample matrices including nasopharyngeal swabs, saliva and blood plasma and increased the sensitivity into the attomole range, up to a 1000-fold increase compared to direct detection in matrix. In addition, a strong positive correlation was observed between the SISCAPA antigen assay and qPCR detection beyond a quantification cycle (Cq) of 30-31, the level where no live virus can be cultured from patients. The automatable “addition only” sample preparation, digestion protocol, peptide enrichment and subsequent reduced dependency upon LC allow analysis of up to 500 samples per day per MS instrument. Importantly, peptide enrichment allowed detection of N protein in a pooled sample containing a single PCR positive sample mixed with 31 PCR negative samples, without loss in sensitivity. MS can easily be multiplexed and we also propose target peptides for Influenza A and B virus detection.

**CONCLUSIONS:** The Cov^2^MS assay described is agnostic with respect to the sample matrix or pooling strategy used for increasing throughput and can be easily multiplexed. Additionally, the assay eliminates interferences due to protein-protein interactions including those caused by anti-virus antibodies. The assay can be adapted to test for many different pathogens and could provide a tool enabling longitudinal epidemiological monitoring of large numbers of pathogens within a population, applied as an early warning system.

## Introduction

The COVID-19 pandemic starkly revealed that humanity is ill-prepared for such global catastrophes. Rising population density, increasing interactions between people and animals in wild habitats and global mobility make humanity increasingly prone to emergent large-scale infections, i.e. future pandemics. It is clear that pandemic readiness needs to be extended, providing tools that allow early warning of threatening pathogens and subsequent rapid development of vaccines and diagnostics. In particular, robust highly sensitive and specific diagnostics allow screening of large populations for monitoring of pathogen load, disease progression and treatment efficacy, perhaps allowing triage of patients when resources are scarce. Although not without their problems, current nucleic acid amplification tests (NAATs) such as reverse transcription - quantitative polymerase chain reaction (RT-qPCR) have been, and will likely remain, a major tool for large-scale screening of individuals. These types of tests work by detecting amplified levels of pathogen-derived nucleic acid and are excellent for testing for exposure to the pathogen. However, there is a need for measuring viral load as a more direct determination of productive virus infection, for monitoring disease progression and treatment and for methods that complement the notoriously sensitive PCR tests. Indeed, the most recent estimation for life virus and thus infectivity in patients was below Cq 31 on the E-gene, effectively showing that higher detections are questionable assets to population testing ^1^.

Several groups have suggested that liquid chromatography coupled to mass spectrometry (LC-MS) might be a method of choice for unequivocal detection of SARS-CoV-2 proteins ^2,3,12,4–11^. In Phase 1 of a community-based effort involving 15 labs and industrial partners, named Cov-MS, we examined the current state-of-the-art for direct LC-MS detection of viral proteins in the most commonly used virus transport media ^2^. We noticed that LC-MS assays for several of the structural SARS-CoV-2 proteins can be developed without having to change sample matrices or standard procedures. Importantly, the different reports on the use of MS essentially detected many of the same SARS-CoV-2 biomarker peptides, irrespective of the model of their LC-MS instruments, or the sample preparation platform or methods used for bioinformatics analysis **(Figure 1)**. In other words, preferentially detected peptides in a preliminary screen turn out to be universally applicable. Thus, we demonstrated that a Phase 1 assay can be developed quickly and without the requirement for clinicians to adopt different sampling procedures.

**Figure 1.**
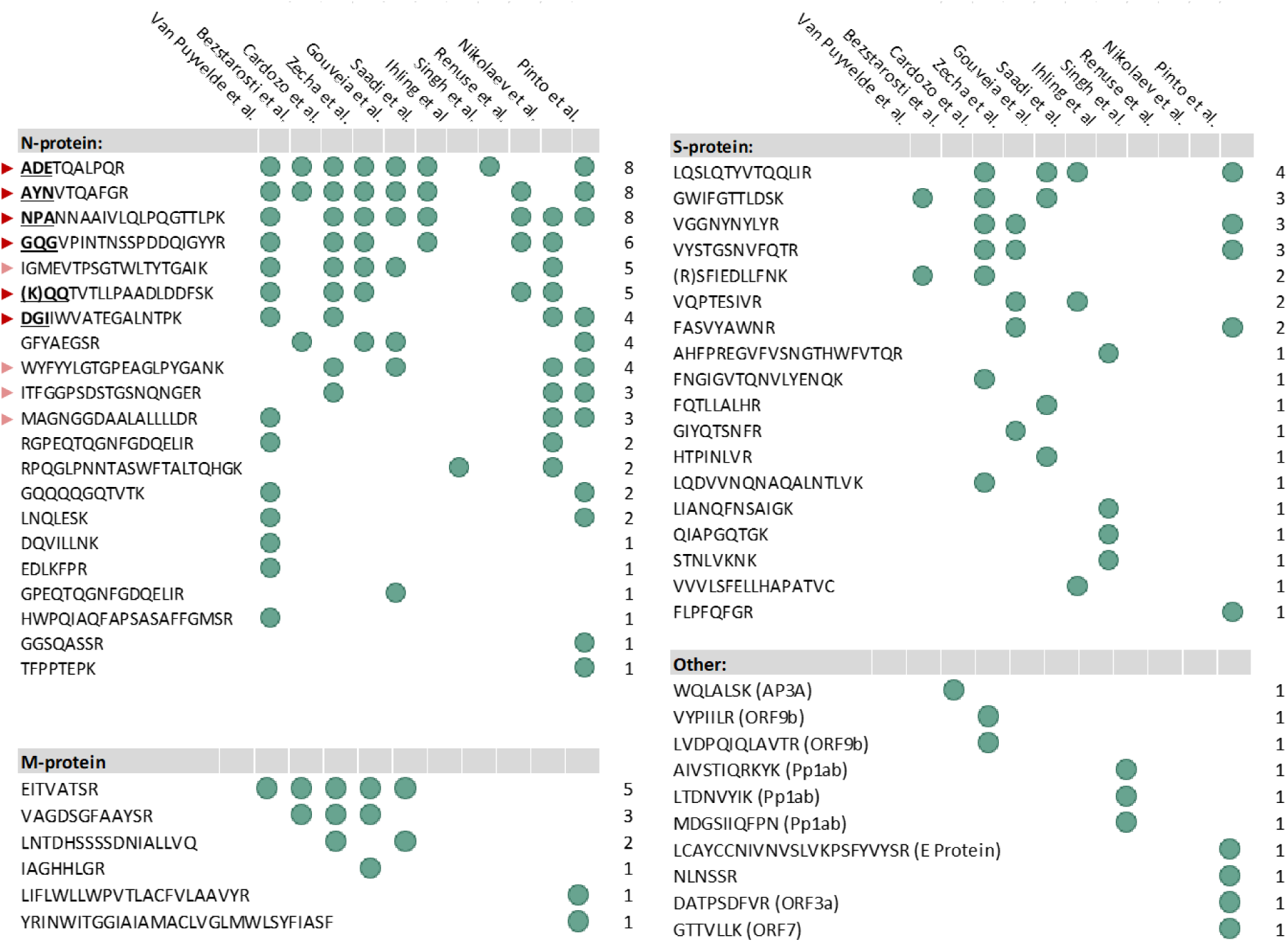
SARS-CoV-2 peptides detected in early MS-based efforts ^2,3,12,4–11^. Red arrowheads indicate peptides targeted for SISCAPA antibody-based peptide enrichment (opaque: polyclonal only). The bold underlined initial three letters depict the peptide abbreviations used throughout the manuscript.

However, there are inherent shortcomings in standard LC-MS methods for peptide identification and quantification. While the latest generation instruments generate enough peptide multiple reaction monitoring (MRM) signal for clinical relevance, the limiting factor is the signal to noise ratio ^13^. It is predominantly the presence of matrix that interferes with and hampers robustness and sensitivity of the Phase 1 Cov-MS assay. In addition to interferences in the matrices, contaminating and potentially interfering molecules are present in most common viral universal transport media that often contain fetal bovine serum (FBS) and thus background proteins. Such proteins contaminate the instrument, limit the amount of sample that can be analyzed (on-column limitations), suppress ionization of analyte peptides, require long chromatography times for peptide separation, and can directly interfere with MS analysis, especially hampering data interpretation.

Here, we build on previous insights and describe the development of a second generation assay, which we named Cov^2^MS, by implementing SISCAPA immuno-MS peptide quantitation technology (13) to reduce or eliminate interferences and to reduce liquid chromatography time, allowing high throughput. SISCAPA peptide enrichment technology has been shown to improve SARS-CoV-2 protein detection considerably ^13,14^. Here we report the full impact of this sample preparation step that enables a more generalized diagnostic method that could readily be deployed in clinical laboratories. We illustrate that the use of peptide immuno-enrichment technology for MS-based SARS-CoV-2 detection essentially addresses the most important issues identified in Phase 1 of the Cov-MS assay. The Cov^2^MS assay can now be applied for the analysis of samples in almost any matrix, i.e. transport medium or biological background, with limited compromise, while increasing its sensitivity into the attomole range, enabling a strong positive correlation with qPCR-based viral RNA levels at least up to Cq 30. The entire workflow is amenable to automation using commercially available liquid handling robots. A single robot can process up to 500 samples in an 8 hour shift. The main factor that limits throughput is the LC-MS time. Still, processing 500 samples per day per instrument is feasible with a cycle time of approximately 2 minutes as presented in this manuscript. Importantly, we have demonstrated that using SISCAPA enrichment, the specimen from one positive patient can be pooled with samples from at least 30 negative patients, without noticeable loss in sensitivity. An exciting observation was that during the development of this updated Cov^2^MS assay, two SARS-CoV-2 variants emerged that spread in the population, including the notorious Delta B.1.617.2 variant-of-concern (VoC). Both variants have mutations in the peptides used in the assay. The mutated forms of the peptides were both enriched using the SISCAPA protocol and were identified by MS, indicating that the Cov^2^MS assay can differentiate between the Delta and other variants simultaneously.

As a future perspective, we propose to establish a candidate target panel of SISCAPA-based-LC-MS assays, which will be able to detect peptides from Influenza A and B viruses at similar or improved sensitivity as SARS-CoV-2. We propose the use of *Infection Proteomics* as a general term for future extensions of the assay. Indeed, next generation tests will have the potential to detect several pathogens simultaneously in most or all transport media or biological media under investigation at sensitivities matching infectivity limits, with considerably higher quantitative accuracies and unequivocal identification of the analyte detected. Especially in light of pandemic readiness, we foresee longitudinal population-wide monitoring of up to a dozen respiratory viruses in pooled patient samples as an early warning system for impending epidemics and pandemics ^13^.

## Materials and Methods

### Materials

Recombinant nucleoprotein (N) of SARS-CoV-2 (2019-nCov), Influenza A (A/Wisconsin/588/2019 – A/Victoria/2570/2019) and Influenza B (B/Phuket/3073/2013) was produced in insect cells with a baculovirus expression system (Sino Biological, Beijing, China).

The SARS-CoV-2 sample preparation protocol was automated using the Andrew Alliance Pipette+ and Shaker+ connected device (Waters Corporation, Milford, MA, USA) both operated via the OneLab platform.

### Virus cultures

#### a) Influenza A (H3N2/2006 - H1N1/2009 Pandemic)

MDCK (Madin–Darby canine kidney) cells were cultured and seeded in complete DMEM medium containing fetal bovine serum (FBS) and non-essential amino acids. The cells were seeded for confluency in a 225 ml Falcon flask at 37°C and 5% CO_2_. Upon reaching confluency, the medium was refreshed with serum-free DMEM containing 4 µg/ml of N-*p*-Tosyl-L-phenylalanine chloromethyl ketone (TPCK) and the adherent cells were infected with mouse-adapted H3N2 (X47) or mouse-adapted H1N1/2009 Pandemic influenza A virus.. One day post infection, cytopathic effects were observed and the supernatant was harvested. The supernatant was centrifuged for 15 mins (4° C; 300 x g) to pellet the cell debris and the clarified supernatant was pooled and frozen at −80°C. A hemagglutination assay was performed which revealed a titre of 2^9^ and 2^4^, respectively, for the X47 and H1N1/2009 pandemic virus-containing supernatant. For this assay, a 2-fold dilution series of the supernatant was made in PBS in a U-bottomed 96 well plate. PBS was used as a negative control. Fifty µl of 1% turkey RBC were added to 50 µl diluted supernatant sample and the mixture was incubated for 1 hr at room temperature, after which RBC agglutination was visually determined.

#### b) Influenza B (Memphis/12/97 – Washington/02/2019 – Phuket/3073/2013)

MDCK cells were cultured, seeded and infected according to the protocol outlined above. Cells were infected with B/Memphis/12/97, B/Washington/02/2019, or B/Phuket/3073/2013 and the cell culture supernatant was harvested and clarified as above, at 3 days post infection, when cytopathic effect was evident. Upon harvest, a hemagglutination assays were performed to confirm viral proliferation. Hemagglutination titers of 2^12^, 2^8^ and 2^7^, respectively, were obtained for the B/Memphis/12/97, B/Washington/02/2019 and B/Phuket/3073/2013 virus preparations.

### Samples

Residual Covid-19 nasopharyngeal patient samples were obtained from the AZ Delta hospital, Roeselare, Belgium with approval of the University Hospital Ghent ethics committee (BC-09263). These samples were analysed at the clinical laboratory of the AZ Delta hospital using the Allplex 2019-nCoV RT-PCR assay from Seegene Inc. as described by De Smet et al ^15^. In short, RNA was extracted from nasopharyngeal swabs using a STARMag 96 × 4 Viral DNA/RNA 200 C Kit (Seegene Technologies, Walnut Creek, CA, USA) on the Hamilton STARlet workstation, followed by real-time PCR using the Allplex SARS-CoV-2 assay. PCR amplification was performed using a CFX96 real-time thermal cycler (Bio-Rad Laboratories) and data were analysed with the SARS-CoV-2 Viewer (Seegene). The presence or absence of SARS-CoV-2 RNA was determined by RT-PCR combined with multiplexed fluorescent probing, in which three different SARS-CoV-2 genes i.e. E-gen (FAM), RdRP (Cal Red 610) and N gene (Quasar 670) and an internal control (HEX) were targeted.

Lyophilised recombinant N protein from Sino Biological was reconstituted to a concentration of 0.1 µg/µL in 100 mM (NH_4_)HCO_3_. A 50,000 amol/µL calibration standard of N was prepared in SARS-CoV-2 negative nasopharyngeal swab pools of different media i.e. 100 mM (NH_4_)HCO_3_), Copan Universal Transport Medium (UTM), Bioer UTM, Sigma Virocult, eSwab, PBS, Plasma, Synthetic saliva (saliva substitute, donated by University of Leicester)- and Patient saliva. A serial dilution was then performed using the SARS-CoV-2 negative nasopharyngeal swab pools to obtain a dilution series with the following concentrations of N: 10,000, 2000, 400, 80, 16, 4, 2 and 0 amol/µL.

An equimolar dilution series of recombinant N protein from Influenza A (Victoria/2570/2019, Influenza B (Phuket/3073/2013) and SARS-CoV-2 (root (L) strain) was generated in 100 mM (NH_4_)HCO_3_. The dilution series contained the following concentrations: 10,000, 2000, 400, 80, 16, 4, 2 and 0 amol/µL.

### Protein extraction and digestion

Each sample was prepared using the same workflow, namely, proteins in 180 µL of sample (for plasma only 60 µL) were precipitated by adding 7 volumes of ice-cold acetone (−20°C). After centrifugation at 16,000 g, at 0° C, the supernatant was discarded and 1 µg of trypsin/lys-C mix (Promega, Madison, WI, USA) in 150 µL 100 mM (NH_4_)HCO_3_ was added. Prior to incubation for 30 minutes at 37° C, to facilitate trypsin digestion, the samples were transferred from Protein LoBind tubes into a 96-well sample collection plate (Waters Corporation). To inhibit further digestion, 50 µL of a 0.22 mg/mL TLCK (Sigma-Aldrich, St. Louis, MO, USA) in 10 mM HCl solution was added to each sample, followed by mixing the plate on the Shaker+ at 1000 rpm for 5 minutes at room temperature. Note that each sample was spiked with 100 fmol of the Cov-MS QconCAT standard (Polyquant, Bad Abbach, Germany) before acetone treatment to precipitate proteins ^16^.

### Peptide selection

Proteotypic SARS-CoV-2 peptides were first identified by *in silico* analysis of the major structural proteins, S, M and N of SARS-CoV-2 and experiments were performed in multiple labs in the consortium to detect the analyte peptides by standard LC-MS protocols ^2^. Different groups detected many of the same proteotypic peptides, irrespective of the instrumentation, sample processing workflow or methods used for bioinformatics analysis. This allowed us to select the “best” peptides (strong, robust MS signals) as surrogates for the viral proteins. We then chose to specifically develop assays for measuring the viral nucleocapsid protein (N) as it is likely to be severely constrained in its structure and less likely to be involved in immunoselection as it is not exposed externally. So far, the N protein encoding gene has exhibited fewer mutated forms than other SARS-CoV-2 structural proteins ^17^. The N protein is also the most abundant of the viral structural proteins with an estimated ∼300-1000 copies per virus particle making it an attractive target for LC-MS-based detection compared to other viral proteins ^18–20^.

### Anti-Peptide Antibodies and Magnetic Bead Immunoadsorbents

Antibodies were raised in rabbits against different proteotypic peptides from the SARS-CoV-2 N protein. Affinity-purified anti-peptide polyclonal antibodies specific for 10 different peptides were prepared and tested in SISCAPA peptide enrichment-MS assays. Of these, 6 polyclonal antibodies yielded excellent results, thus triggering the derivation of rabbit anti-peptide monoclonal antibodies (RabMAbs) from the same rabbits. The RabmAbs were screened by proprietary methods to characterize their ability to bind peptides from complex tryptic digests of plasma, saliva and nasopharyngeal swab materials and to select those that enrich peptides without detectable interferences. This process allows selection of highly specific, high affinity anti-peptide antibodies capable of binding low abundance peptides from solution and retaining them through extensive washing steps designed to minimize non-specific background. The selected antibodies were produced as recombinant proteins and all have sub-nanomolar affinities and, more specifically, slow off rates. Antibodies specific for six peptides (ADETQALPQR, AYNVTQAFGR, DGIIWVATEGALNTPK, NPANNAAIVLQLPQGTTLPK, GQGVPINTNSSPDDQIGYYR and KQQTVTLLPAADLDDFSK) were covalently coupled using dimethyl pimelimidate to protein G magnetic beads (Life Technologies) in 1 x PBS with 0.03 % CHAPS and stored at 4-8° C.

### Peptide enrichment

Antibody-coupled magnetic bead immunoadsorbents were resuspended fully by Vortex mixing. Equal volumes of the six bead suspensions were mixed and 60 µL of the mixture were added to the trypsin-digested samples, once tryptic digestion activity had been neutralized. Plates were put on the Shaker+ and first shaken at 1400 rpm for 3 minutes to ensure that beads were fully resuspended prior to a one-hour incubation at 1100 rpm at room temperature. After incubation, the plates were placed on a custom-made magnet array (SISCAPA Assay Technologies) for 1 minute and once the beads had been drawn to the sides of each well, the supernatant (approximately 260 µL) was removed. The beads were then washed by addition of 150 µl wash buffer (0.03 % CHAPS, 1 x PBS) to each sample followed by resuspending the beads by shaking the plates at 1400 rpm for 1 minute. The sample plates were then again placed on the magnet array and the supernatant was removed. The washing step was performed a second time. Subsequently, the beads were resuspended in 50 µl elution buffer (1 % formic acid, 0.03 % CHAPS) and mixed at 1400 rpm for 5 min at room temperature. Finally, after placing the plates on the magnetic plate, the eluents containing the eluted peptides were transferred to a QuanRecovery 96-well plate (Waters Corporation) for LC-MS analysis.

### LC-MS Detection and Quantification

LC separation was performed on an ACQUITY UPLC I-Class FTN system, with Binary Solvent Manager and column manager with column selection valves (Waters Corporation). Ten microliters of the enriched sample were injected onto an ACQUITY Premier Peptide BEH C18 column (2.1 mm x 30 (or 50) mm, 1.7 µm, 300 Å) column (Waters Corporation). Peptide separation was performed using a gradient elution of mobile phase A containing LC-MS grade de-ionised water with 0.1 % (v/v) formic acid and mobile phase B containing LC-MS grade acetonitrile with 0.1% (v/v) formic acid. For the 1-minute run, gradient elution was performed at 1 mL/min with initial inlet conditions at 7 % B, increasing to 40 % B over 0.3 min, followed by a column wash at 90 % B for 0.25 min and a return to initial conditions at 7 % B. The total run time was 0.8 min. For the 2-minute min run, gradient elution was performed at 0.8 mL/min with initial inlet conditions at 5 % B, increasing to 15 % B from 0.15 to 0.35 min and at a steady state for 0.25 min. Subsequently, over 0.4 min, gradient B was increased to 25%, followed by a column wash at 90 % B for 0.25 min and returning to initial conditions at 5 % B. The total run time was 1.8 min. For the 8-minute run, gradient elution was performed at 0.6 mL/min with initial inlet conditions at 5 % B, increasing to 33 % B over 5.5 min, followed by a column wash at 90 % B for 1.4 min and a return to initial conditions at 5 % B. The total run time was 8 min.

A Xevo TQ-XS tandem MS (Waters Corporation, Wilmslow, UK), operating in positive electrospray ionization, was used for the detection and quantification of the peptides. The instrument conditions were as follows: capillary voltage 0.5 kV, source temperature 150° C, desolvation temperature 600° C, cone gas flow 150 L/h, and desolvation gas flow 1000 L/h. The MS was calibrated at unit mass resolution for MS1 and MS2. Endogenous and the corresponding stable isotope labelled peptides were detected using MRM acquisition with experimental details described in Supplemental Table 1.

Skyline (version 21.1) was used to process the raw LC-MS data using a template file containing the six target peptides. Peak integration boundaries were automatically set on the heavy standard and manually reviewed before exporting a report containing the peptide modified sequence, transition, area and height among others.

## Results

### Development and preliminary validation of SISCAPA anti-peptide antibodies

Throughout the SARS-CoV-2 pandemic, clinical laboratories have switched between providers of nasopharyngeal swabs and transport media because of fluctuations in the supply chain. All of these were validated for qPCR compatibility. However, we have shown that transport media can heavily impact the detectability of viral proteins using MS ^2^. Therefore, to improve the detection and measurement of viral proteins by LC-MS it was deemed desirable to remove interferences as much as possible. To do this, SISCAPA-compatible high affinity anti-peptide antibodies were produced against a selection of proteotypic, surrogate N protein peptides **(Figure 1, red arrowheads)**. These peptides were originally also incorporated into the Cov-MS QconCAT heavy internal standard (PolyQuant, Bad Abbach, Germany), which was also used here throughout the assay development process ^16^.

Several other groups worldwide have assessed the detectability of SARS-CoV-2 peptides using LC-MS. Encouragingly, many of the targeted peptides proved universally applicable **(Figure 1)**. The N protein is the most favorable target protein, in part because of its high copy number in the viral particle, with roughly 30-35 ribonucleoprotein decamers or >300 N protein molecules per particle ^20^. N protein is also likely to be severely constrained in its structure and less likely to be involved in immunoselection, as it is not exposed externally.

Polyclonal antibodies were first raised in rabbits by immunization with the selected proteotypic, tryptic peptides. Affinity-purified polyclonal antibodies were used to further study the ten SISCAPA peptide assays in the context of appropriate sample matrices. Subsequently, six targets were chosen to proceed to the second phase of the development where rabbit monoclonal antibody reagents were produced at scale. An in-house comparison of the performance of polyclonal and monoclonal antibodies for two of the peptides is shown in **Figure 2A**. The increased performance of monoclonal antibodies is especially pronounced at lower concentrations, effectively increasing the sensitivity of the assay where it is most needed.

**Figure 2:**
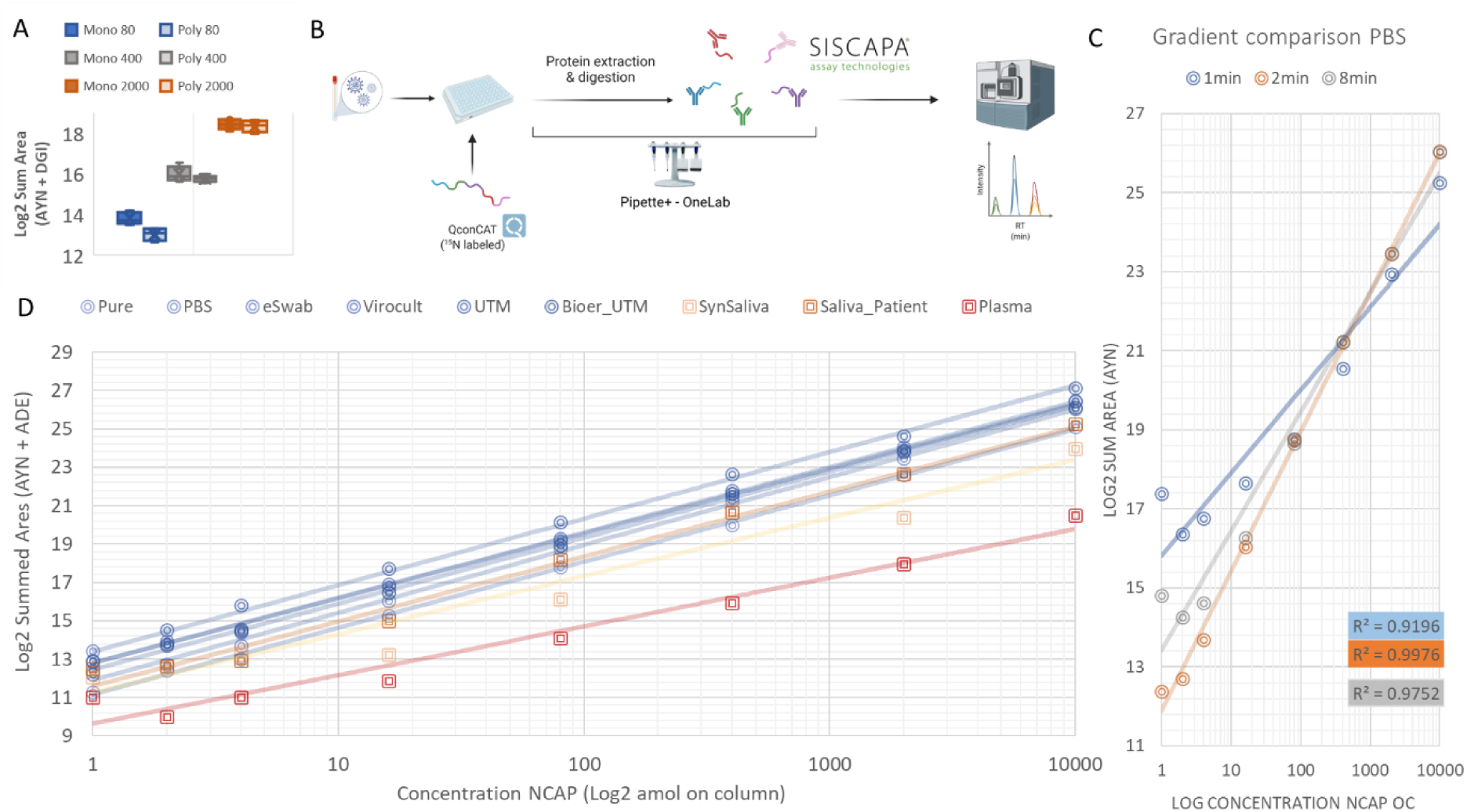
Validation of the peptide enrichment protocol using SISCAPA technology. **(A)** Intensity comparison (LogSum area under the curve) between polyclonal and monoclonal antibodies against two target peptides (AYN and DGI) spiked in three different concentrations (80-400-2000 amol/µL) in Bioer UTM background. **(B)** Schematic representation of the SISCAPA workflow. **(C)** Comparison of different gradient lengths and their linearity for detecting the AYN peptide. **(D)** Linearity of detection of recombinant N protein spiked in a dilution series in different matrices. 100 mM (NH_4_)HCO_3_, PBS, Copan eSwab, Sigma Virocult, Copan UTM, Bioer UTM, Synthetic saliva, Saliva patient and Plasma.

### Automation and purification lead to higher sample throughput

The “addition-only” SISCAPA protocol relies on the use of magnetic bead immunoadsorbents for target peptide purification and can therefore be automated to both increase the sample throughput and to reduce technical variation, i.e. increase quantitative accuracy. Therefore, we optimized a protocol using the programmable Pipette+ system which in time can be transferred to liquid handling robots. **Figure 2B** shows this setup and **Supplemental Figure 1** provides a snippet from the OneLab interface used to intuitively program the Pipette+ protocol, which is available together with the raw data on Panorama Public. While still under development at the time of data collection, a complete sample preparation kit from Waters is now available ^23^, which is expected to further standardize the assay and facilitate automation. Note that in the current work, we used acetone precipitation of proteins and re-solubilization in (NH_4_)HCO_3_ instead of the RapiGest surfactant supplied in the kit.

After removal of non-specific matrix molecules, as achieved by the peptide enrichment method, the LC gradient can be shortened without the risk of increasing interferences, providing the additional benefit of narrowing the chromatographic peaks, providing increased detectability. This should however be balanced against the number of MRM transitions and required points-per-peak to maintain sufficient ion statistics and thus quantitative precision. Therefore, we compared the original Cov-MS gradient of 8 minutes with a gradient of 2 minutes for all peptides, and a 1-minute gradient for only 2 peptides (AYN and KQQ) and assessed the linearity of a dilution series in PBS matrix. **Figure 2C** shows the response for AYN in Log2 transformed intensity (LogInt). **Supplemental Figure 2** summarizes the individual responses of the six different peptides indicated with red arrowheads in **Figure 1**. Overall, a 2-minute gradient demonstrated the highest linearity, with the ADE and AYN peptides performing best in terms of MRM sensitivity, especially in the low intensity range close to the lower limit of quantification. While the KQQ peptide showed the best linearity using the 8-minute LC gradient, manual inspection of the data showed that the apparent higher signal within the low intensity range for the shorter gradients was caused by carry-over effect in-between dilution series.

Notably, using a two-minute gradient, a total of 500 patients per instrument per day could potentially be analysed. While such patient sample batch sizes are not yet available, 600 samples were run in less than 96 h on two separate occasions during the work reported here, including dilution series comprising a total of nine different matrices. No decline in instrument performance was apparent. In summary, a 2-minute gradient provided the highest linearity, detectability and throughput, and was selected for subsequent analyses.

### Mitigating matrix effects

The assessment of matrix effects that can affect LC-MS is an essential part of method development, especially with respect to addressing any issues related to future viral transport medium availability. Additionally, enabling direct peptide detection in a variety of biological matrices significantly broadens the applicability of the method. Therefore, we assessed the linearity of response for six N protein peptides in a dilution series in (i) six different (viral transport) media and (ii) two different biological matrices, saliva and plasma, as well as a synthetic surrogate for saliva. **Supplemental Figure 3** shows the light/heavy ratio of all peptides in the different matrices. Three peptides showed a high dynamic response with low variability, i.e. AYN, ADE and KQQ.

Two peptides (ADE and AYN) were selected for further work based on their stability and linearity using 2-minute gradient in all the transport media tested. **Figure 2D** shows their signal intensity (area sum MRM transitions) in these different media. Note that for the combined sum of the intensities for these two peptides, the blank signal is lower than that at 72 amol on-column in all transport media. This is in line with our previous estimations, where a theoretical detection limit of 40 amol was proposed in samples without matrix ^2^. This in turn demonstrates the efficiency of the enrichment strategy. In fact, for Universal Transport Medium (UTM), this dilution series implies at least a 100-fold more sensitive detection compared to the Phase 1 Cov-MS. Note that the previous UTM detection limit estimation was heavily compromised by interferences and that the peptide selection made by a machine learning based approach was matrix specific ^2^. The 100-fold increase in signal is therefore a conservative estimation, as will be illustrated below.

The new assay also performs very robustly when using saliva and plasma. Thus, a variety of sample matrices could potentially be used in the future if the viral load is adequately high in such biological fluids ^24,25^. Equally important, the detection of viral peptides in plasma creates the possibility for direct detection of viral load in blood, in turn enabling the assessment of disease status, clinical prognostic value and treatment monitoring. It will be interesting to test for its utility for monitoring of long COVID, perhaps using addition biomarkers for inflammation or immune responses in a multiplexed analysis ^26^.

### A comparison between RT-qPCR and SISCAPA-LC-MS performed on patient samples

While five monoclonal antibody reagents were used on the 233 patient sample batch, we first assessed the performance of the AYN peptide as suggested by Hober et al. ^14^. A 2-minute gradient was used and three different viral transport media were included, i.e. PBS, Bioer UTM and Bioer VIM (Viral Inactivation Medium). Note that in order to define the sensitivity and specificity for MS analysis, high patient numbers and a ground truth are required, e.g. by using true negative patients sampled before the pandemic. Such samples were unavailable to us. Therefore, a binary comparison to qPCR (positive and negative) should rather be expressed in percent positive agreement (PPA) and percent negative agreement (PNA) ^14,27^. **Figure 3A** depicts these numbers when an initial summed MRM intensity of AYN of 26 LogInt is used to define positive patients for MS and < Cq 40 is used for the qPCR threshold (122 qPCR-positive and 111 qPCR-negative patients). As suggested by Hober et al., we also plotted these numbers against a qPCR positivity threshold of Cq 30 - leaving out the patients between Cq 30 and 40. This clearly illustrates how well qPCR and MS agree, especially up to Cq 30, with 96.2% PPA and 98.2% NPA, respectively ^14^.

**Figure 3.**
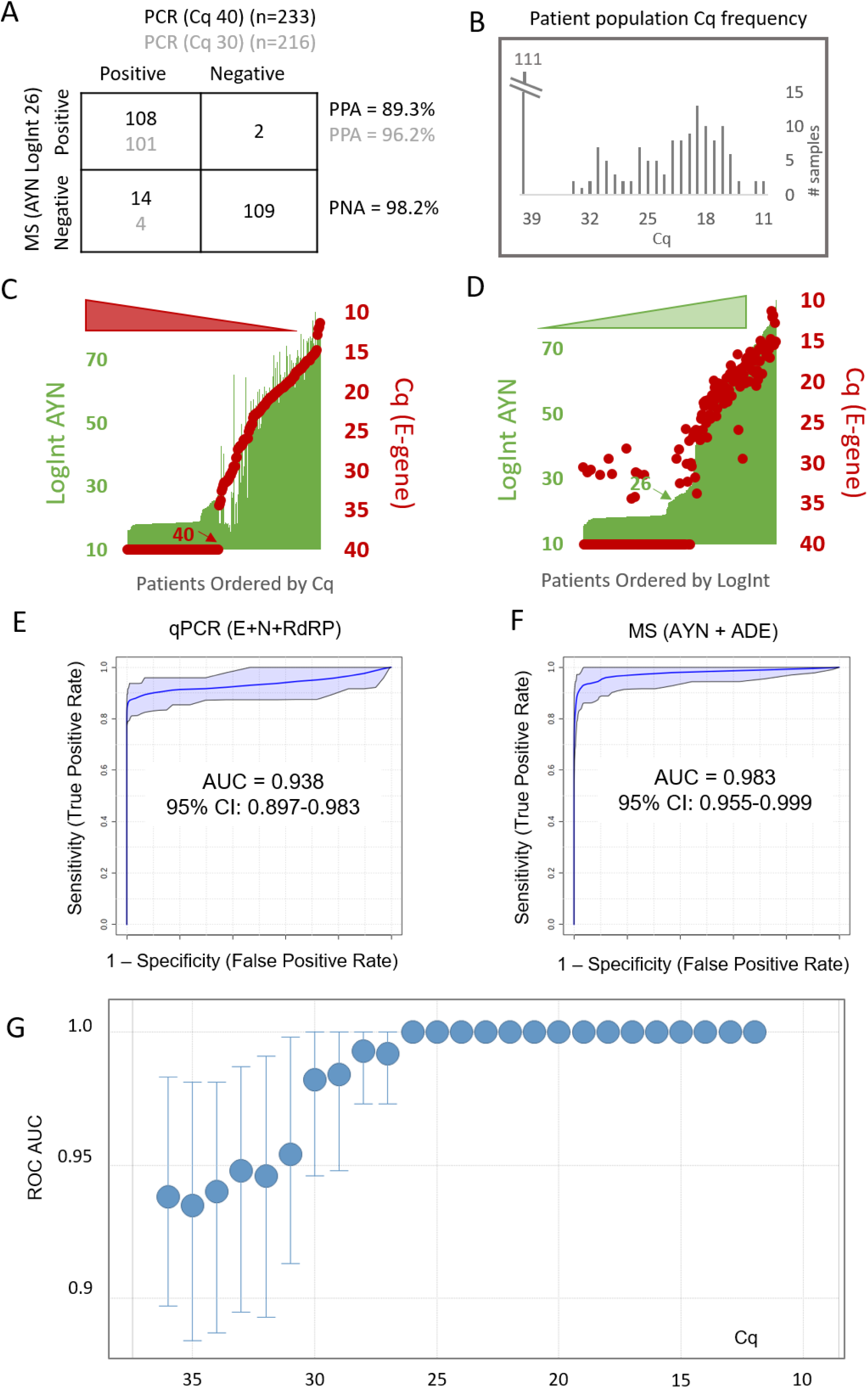
A comparison between RT-qPCR and SISCAPA-LC-MS performed on 233 patient samples. **(A)** A patient sample batch in different media displays a high percent positive (PPA = TP/(TP+FN)) and negative agreement (PNA = TN/(TN+FP)) between qPCR (Cq) and MS (LogInt), especially below Cq 30 (grey numbers). **(B)** The distribution of the patient population over their measured E-gene Cq values. The frequency of patients will greatly impact the PPA and PNA at different Cq thresholds. **(C and D)** Secondary axis plots of the raw measurements of E-gene Cq (red dots) and AYN Logarithmically transformed MS intensities (LogInt) (green bars) for patients sorted from high to low Cq **(C)** and low to high LogInt **(D)**. A strong linear correlation illustrates the level of agreement between both tests. **(E and F)** ROC curves with true positives defined by either qPCR **(E)** or MS **(F)**. AUC: area under the curve. **(G)** ROC AUCs for each Cq value separately. Up to Cq 26, there is perfect agreement (AUC = 1). Above Cq 30, a noticeable drop-off to AUC of 0.95 can be seen, suggesting that from here on both diagnostic tests start to disagree. Error bars indicate the 95% confidence interval (CI).

However, there are several concerns with this representation. First, both tests report a continuous measure rather than a binary outcome and a threshold needs to be chosen to define “positive” and “negative”. This is arguably not trivial for either test. In fact, the positivity threshold for qPCR varies greatly between assays and is currently set to Cq 33 for the applied test in the hospital providing the patient samples for this study and this should probably be set to Cq 31 in light of recent insights on infectivity ^1^. Second, the numbers reported in the matrix are a direct function of the Cq distribution in the patient population tested. **Figure 3B** illustrates that shifting e.g. the positivity threshold of qPCR from Cq 40 to Cq 35 would not impact PPA or PNA because there are no patients that sample that region. Third, qPCR is not well suited to define the amount of RNA in copies/ml, with measurements sometimes differing > 1000-fold between laboratories ^28^. MS on the other hand is considered a quantitative and accurate analytical tool, an asset typically not attributed to other protein detection technologies, such as laminar flow antigen tests ^29^. In other words, the Cq reported for a patient can vary greatly and thus defining qPCR as the ground truth or golden standard does not objectify the comparison. Finally, there is a potential underlying biological reason why RNA and protein do not completely correlate, i.e. the stage of infection ^30,31^. Indeed, while RNA and protein levels will most probably rise in parallel at the onset of infection, it is known that residual RNA can still be detected over a month following infection when the disease symptoms are no longer apparent ^1,31,32^.

Therefore, the raw results from both tests are first depicted on two secondary axis plots (**Figure 3C and D**). Each patient (x-axis) is represented by its two respective measurements, i.e. LogInt of the AYN peptide (green bars, left axis) and the Cq value for the E-gene (red dots, right axis). As reported earlier, the LogInt of positive patients strongly correlates linearly (R^2^ = 0.86) to the Cq value, which is an exponential metric depicting the number of doublings required for detection ^2,14^. In both plots, patients were sorted from low to high virus measurement, i.e. from high to low Cq in **Figure 3C** and from low to high LogInt in **Figure 3D**. While only two patients with Cq 40 had a LogInt for AYN>26, evenly spread patients have Cq values of 28-35 below this MS intensity threshold, all the way down to the patient with the lowest value as measured by MS. These samples are either false positives by qPCR or false negatives by MS as depicted in **Figure 3A**. Alternatively, this raises the possibility that these patients were in an early or late stage of infection ^31^.

Next, we calculated receiver operating characteristics (ROC) using multivariate ROC curve analysis based on linear support vector machine (SVM) using all five peptides **(Figure 3 E and F)**. Classes were defined based on qPCR diagnosis together with log summed MRM area values of the investigated peptide features. **Supplemental Figure 4** shows the contribution of the different genes (qPCR) and peptides (MS) to diagnosis as expressed in Selected Frequency % (SF%). A t-test was used to coordinately assess the significance of the difference of each of these measurements between positive and negative patients defined by the other test. For MS, a clear distinction was seen in SF % between AYN (SF % = 1.0; p = 1.1 e-51) and ADE (SF % = 1.0; p = 1.3 e-37) on the one hand and the three other peptides in the assay on the other, i.e. KQQ (SF % = 0.5; p = 1.4 e-30), DGI (SF % = 0.3; p = 8.5 e-28) and NPA (SF % = 0.25; p = 2.8 e-26). Note that all peptides have a low t-test statistic and thus do differ significantly between positive and negative patients (the GQG peptide was not immuno-purified from the patient samples), yet, peptides ADE and AYN again performed best. For the qPCR, the order of performance was E gene (SF % = 0.9; p = 4.2 e-31), RdRp (SF % =0.7; p = 1.4 e-31) and then N gene (SF% =0.35; p = 1.0 e-31) for best classifying patients diagnosed by MS. Notably, the E-gene was recently proposed to correlate best to infectivity ^33^. Whether this correlation also implies that MS correlates well with infectivity remains to be determined.

**Figure 3 E and F** show the ROC curves with qPCR and MS respectively defining the ‘ground truth’ at the clinical diagnosis level of Cq > 36 or the summed LogInt for the AYN + ADE peptides at 26.6, which was inferred from the median and median absolute deviation values of the patient distribution shown in the B panel. It is clear from the ROC area under the curves (AUC) and their confidence intervals that both tests largely agree. In a final effort to realize a threshold for MS in light of qPCR, we plotted all the ROC AUCs for each qPCR threshold **(Figure 3G)**. This shows perfect agreement (AUC = 1) up to Cq 26 and only above Cq 30, a noticeably drop-off to AUC of 0.95 can be seen, suggesting that from here on both diagnostic tests start to disagree slightly. Still, it is important to take the patient population distribution into account when interpreting these thresholds in the higher Cq region **(Figure 3B)**.

**Supplemental Figure 4C** shows the linear correlation between LogInt AYN and Cq separately for the different media in the sample batch, illustrating how the transport medium has only minimal effect after peptide enrichment. Importantly, the initial Cov-MS assay without matrix removal lost linearity around Cq 20-21 for UTM samples with considerable MRM signal interference ^2^. As Cov^2^MS can now measure positive patients up to Cq 30 in UTM, this implies close to a 1000-fold increase in assay sensitivity for patient samples in UTM matrix. Importantly, this patient data also very strongly resembles the results obtained by Hober et al., showing effective inter-laboratory roll-out ^14^.

We anticipate that the assay could hold potential up to Cq 33, because (i) one patient with Cq 32 was detected with considerable MS signal, (ii) The AUC of the ROC curves is still > 0.9 up to the least positive patient in the batch (Ct = 34), (iii) Cq values are known to vary significantly (for lower viral loads) ^28^, (iv) Patients above Cq 30 tend to be rarer than highly positive ones, biasing the resolution in the LOD area, as also seen in the study of Hober et al., (v) the Cov^2^MS assay can theoretically be conducted on larger volumes of sample since it includes an enrichment step, (vi) reducing the LC flow rate to 5 µl/min using 300 µm ID columns, whilst maintaining on-column load, would further increase sensitivity, and (vii) a better metric or method than a regular LogInt threshold could be developed to classify samples. Note that above Cq 30, a retest or physical diagnosis by a physician is usually required, even for qPCR and using a probabilistic score would therefore greatly facilitate the triage of patients with such viral loads. Irrespectively, culturable virus can no longer be detected in specimens with an E gene CT above 31, and a new primer-probe set was recently developed to make sgE undetectable above this value, effectively avoiding quarantine and isolation decisions that do not impact the spread of the virus ^1^.

### Detection of mutations and Variants of Concern

When evaluated using combined peptides ADE + AYN, several patients were significant outliers from the linear correlation with the Cq value **(Figure 4A)**. Yet, they behaved more coherently when only the AYN LogInt signal was plotted. Upon detailed inspection of the most positive sample (Cq 11), there was no signal for the ADE peptide, while the heavy standard peptide from the QconCAT was measured as expected **(Figure 4A insets)**. This indicates that this loss in signal cannot be attributed to sample preparation issues ^2^. Since SAT developed anti-peptide antibodies against peptide sequences from genetically stable regions, variant screening is hampered. Still, in a multiplexed assay a sudden absence of signal from one specific peptide, while maintaining the heavy signal as well as the other peptides, could suggest a mutation in the target sequence **(Figure 4A)**. The loss of signal can be attributed to two complementary phenomena: (i) the MRM assay is not measuring the correct transitions at the altered retention time, irrespective of (ii) whether or not the peptide is still captured by the antibody reagent. To assess the latter, the enriched Cq 11 patient sample highlighted in **Figure 4A** was re-analyzed using discovery Data-Dependent Acquisition (DDA) on a high resolution TripleTOF 6600+ System (SCIEX, Concord, ON, Canada). Manual inspection of the data showed that the targeted peptide was still present in the sample (it was captured by the antibody), yet the N-terminal alanine (A) in the peptide stretch was mutated to a threonine (T) (A376T) **(Figure 4B)**. Note that not a single peptide other than the SISCAPA targets could be detected or identified in these samples using DDA, illustrating the selectivity of the method and thus the purity of the Cov^2^MS peptide samples presented for LC-MS analysis. As the peptide was still immuno-purified, a simple switch of acquisition parameters was sufficient to detect it beyond any doubt using MRM upon re-injection **(Figure 4C)**. To verify the validity of this mutation, we tracked the GISAID (https://www.gisaid.org/) database and found that this particular variant was briefly circulating in Belgium around the time of sample collection **(Figure 4D)**. Note that if the peptide had not been enriched by the SISCAPA workflow, the MRM assay would only be capable of detecting the absence of signal. In turn however, this inspired us to verify if we could enrich and detect the neighboring T377Y mutation known from the Delta B.1.617.2 variant, which arose in Belgium not long after **(Figure 4D)**. Indeed, this peptide too could be identified using high resolution MS **(Figure 4E)** and was detected by a small adaptation in the transitions of the MRM analysis **(Figure 4F)**, as also seen by others ^34^. The mutation had also induced a small retention time shift.

**Figure 4.**
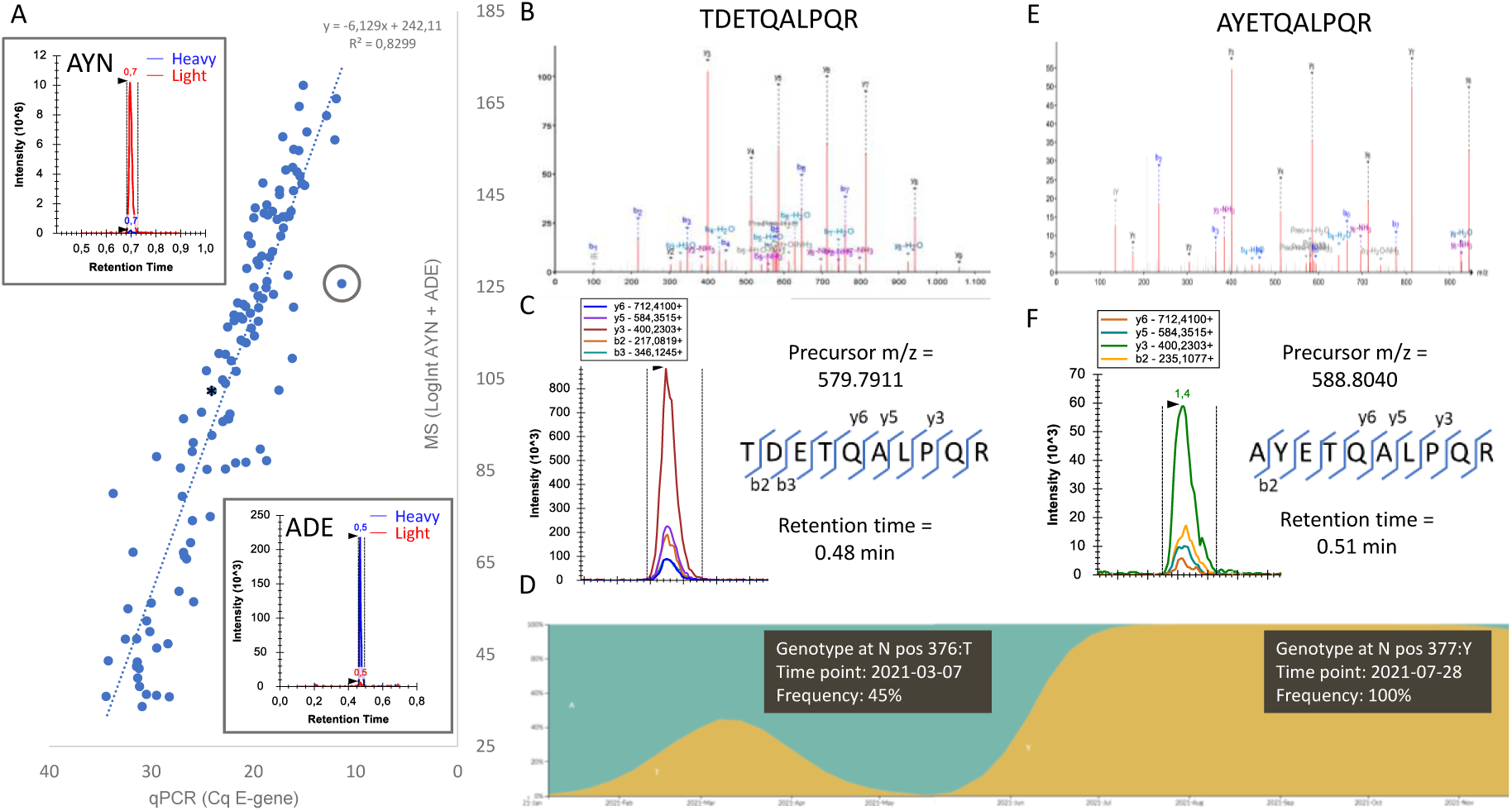
Variant screening. **(A)** Several patients had a summed ADE + AYN LogInt that was significantly lower than what was expected based on the linear correlation with the Cq value. Manual inspection of one patient (with Cq 11, circled in grey), showed how only the light signal of the ADE peptide and not the one from the AYN peptide had disappeared (insets). **(B)** When this patient sample was acquired in discovery DDA, a fragment spectrum was found that could be annotated as TDETQALPQR (A376T). **(C)** The same sample was then re-acquired in MRM, this time targeting the mutated peptide by precursor mass and by two b-ions that contain the mutation; a clear signal could be picked up. Adding these to the assay now allows detecting the mutation in all patients in the batch. **(D)** By checking the GISAID database ^35^ the frequency of this mutation in Belgium showed that this variant was circulating around the time the samples were taken. Not long after, the Delta variant, which contains the (D377Y) mutation, completely replaced the other variants. The figure is composed of two consecutive screenshots. **(E)** Therefore, a similar approach was applied to specifically identify a biomarker peptide for the Delta VoC and the resulting high resolution MSMS spectrum is shown. **(F)** This D377Y mutation was still immuno-enriched by the SISCAPA antibody reagent (somewhat less efficiently) and again the target can easily be added to the MRM, albeit at a slightly shifted retention time.

In conclusion, multiplexing and use of a stable isotope labelled internal standard together allow the detection or absence of signal for a single peptide, indicative of a mutation. If the peptide is still sufficiently enriched by the antibody, it can be readily detected by means of MRM analysis using alternative transition and retention time parameters within the same sample upon the next injection. This emphasizes the importance of targeting a minimum of at least two peptides at all times, in order to avoid positive patients evading detection.

### Peptide immuno-affinity enrichment enables efficient sample pooling

Because of the strongly fluctuating positivity rates of testing throughout a pandemic, it has been proposed to use patient sample pooling to increase throughput and reduce reagent usage for qPCR ^36^. Up to 1/32 pools can be theoretically beneficial for qPCR, depending on the positivity rate and thus the phase of the pandemic. However, for qPCR this leads to a tradeoff in sensitivity since every dilution step reduces the detection by one Cq value, meaning that for a 1/32 dilution experiment, five Cq values in sensitivity need to be sacrificed, implying that e.g. a single positive patient of Cq 30 might remain unnoticed in such a pooled sample. In contrast, in this second generation Cov^2^MS assay, that measures viral protein, the tryptic peptide biomarkers are enriched through antibody binding on magnetic beads. This essentially means that the peptides are extracted from the buffer, theoretically making the assay insensitive to dilution and thus pooling ^13^. Note that we opted to digest the patient samples first and pool fractions of these at the peptide level, as a positive pool always requires re-analysis of the separate patients to pinpoint the positives.

Indeed, for all patients tested, the summed LogInt of ADE + AYN remained stable irrespective of the dilution, i.e. ½, ¼, 1/8, 1/16 and 1/32 dilution with a mixture of negative patient samples **(Figure 5A). Figure 5B** depicts the raw signal of a patient with Cq 26. Note that only one fifth of this patient sample (10 µL out of 50 µL) was used to enable making each of the five different dilutions needed out of this sample. This is not necessarily done in a true world setting wherein e.g. 40 µl can be used. In other words, this is equivalent to using a patient of two Cq higher qPCR results. Indeed, the raw data from all five dilutions in Figure 5C show a lower signal than the undiluted sample for that reason, yet the signal does not decrease according to dilution. This effectively proves that peptide immuno-affinity enrichment is insensitive to pooling over the full range of detectable viral loads.

**Figure 5.**
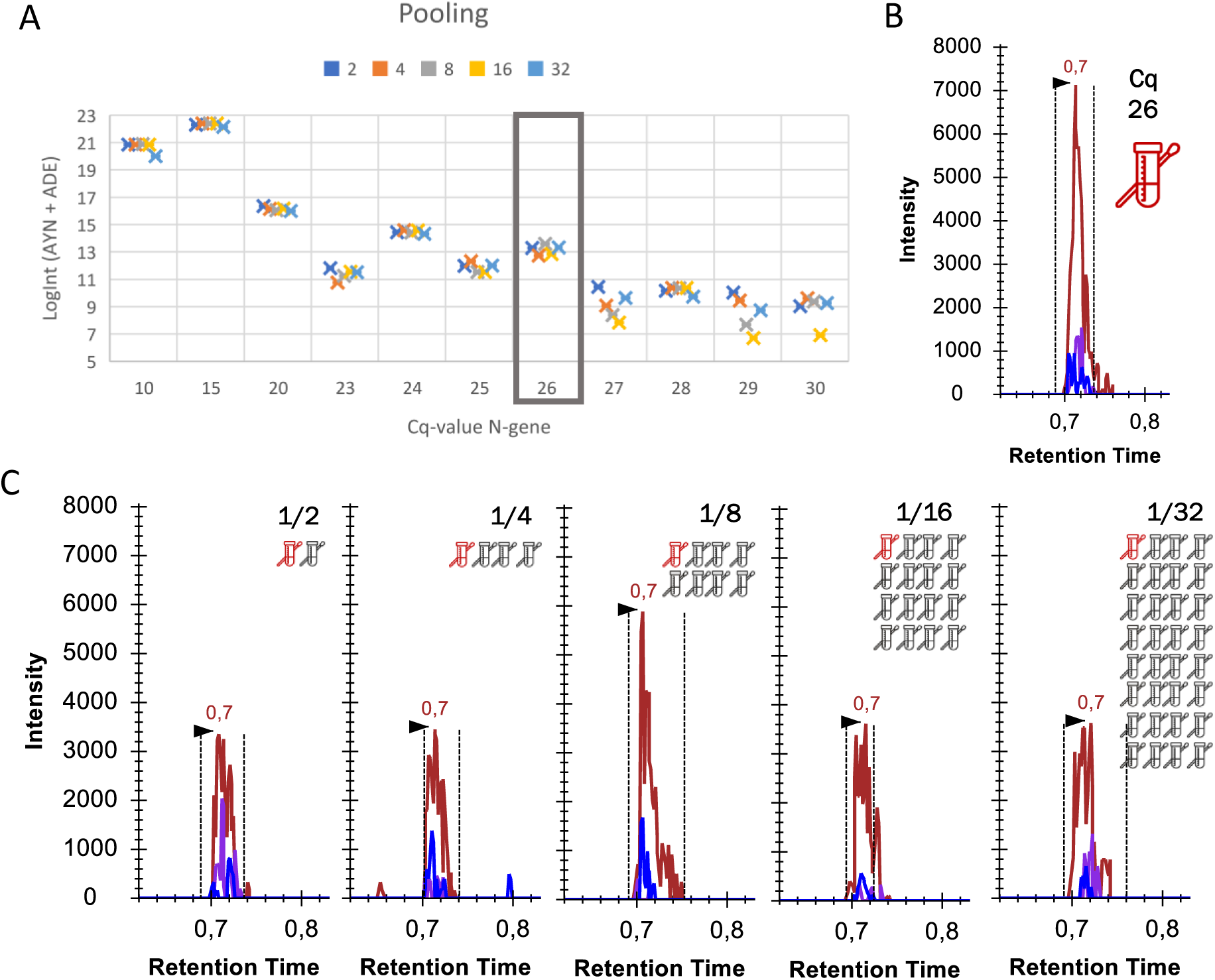
SISCAPA peptide immuno-affinity enrichment is insensitive to patient pooling. **(A)** 10 µl out of 50 µl of the peptide digest of positive patient samples with different Cq values were diluted with q-PCR-confirmed negative patient samples in five different ratios (1/2, 1/4, 1/8, 1/16 and 1/32), with each dilution corresponding to a loss of one Cq-value. However, by using SISCAPA peptide immuno-affinity enrichment, LC-MS is insensitive to the dilution effect, hence a similar signal intensity is achieved for each dilution. **(B)** One positive (red) patient with Cq 26 (boxed in (A)) was manually inspected. The initial measurement of 50 µl sample resulted in a signal for AYN of 7000. **(C)** When another 50 µl of digest from this patient were spilt into five and diluted ½, ¼, 1/8, 1/16 and 1/32 with a digest of mixture of negative patients, the signal did not decline accordingly, effectively showing how the SISCAPA workflow is insensitive to pooling.

### The future of infection proteomics

Because of the urgency, most of the previous and current work centered around detecting SARS-CoV-2 in patient samples. However, the approach used can be expanded to include a wider variety of infectious pathogens since an LC-MS instrumental setup simply measures two key properties of an analyte: its hydrophobicity (through its retention time) and its (fragment) masses (through *m/z* or MRM detection). Therefore, including other pathogens into a multiplexed array can be done rapidly and without compromise, with the gradient time and MS duty cycle being the only limiting factors. Still, it can be estimated that a 2-minute gradient can potentially harbor a sufficient number of transitions to monitor up to a dozen pathogens in a single run. We suggest targeting virus nucleoproteins as a first approach because they interact with the virus nucleic acid (providing the linear correlation with qPCR) and are likely to be structurally highly constrained and thus less likely to accumulate mutations. In addition, mutated forms of nucleoproteins are not likely to be selected for because they are less exposed to the immune system of the host, although antigenic drift on viral nucleoproteins due to recognition by cytotoxic T cells should be considered for certain pathogens ^37^.

As a proof of concept for multiplexed pathogen detection, we repeated the workflow of Phase 1 of Cov-MS to detect peptide biomarkers for Influenza A (H1N1/2009 and H3N2/X47) and Influenza B (B/Memphis/12/97, B/Washington/02/2019 and B/Phuket/3073/2013) virus strains, using viral cultures according to the workflow described in ^2^. After detecting the best ionizing N protein peptides, i.e., highest MRM intensity, we filtered them based on mutational stability, i.e. variant coverage frequency, which was found to be greater than 99%, derived from the *in silico* proteolytic analysis of the 100% UniRef N protein sequences from January 1, 2010 to June 30, 2021 obtained from UniProt Knowledgebase. This led to the consensus targets depicted in **Figure 6**. Interestingly, when measuring an equimolar mixture of recombinant N protein from SARS-CoV-2, Influenza A and Influenza B, the influenza peptides generated two-to three-fold increased signal compared to the peptides presented in the Cov-MS and Cov^2^MS assays. We therefore anticipate that the translation of the assay to other viruses will at least yield comparable or better results. In conclusion, we envision a single multiplexed assay that can be used to detect any of these respiratory pathogens in patient samples while simultaneously differentiating between them, thus significantly lowering the cost of analysis.

**Figure 6.**
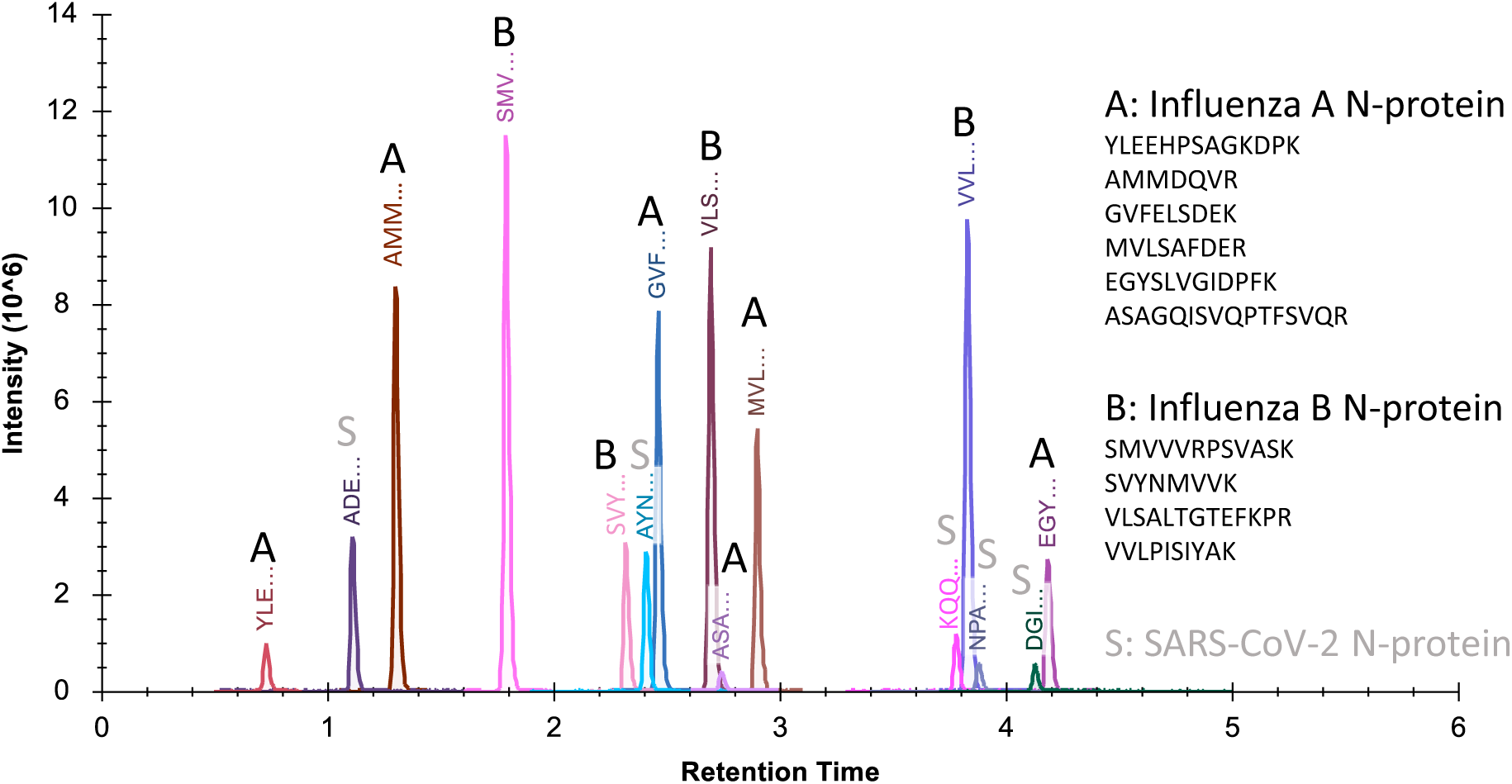
Preliminary data illustrating multiplexed analysis for SARS-CoV-2, Influenza A and Influenza B. The Phase 1 workflow described for the previously published Cov-MS work was applied to viral cultures of different Influenza A (H1N1/2009 and H3N2/X47) and Influenza B (Memphis, Phuket and Washington) strains. Proteotypic peptides specific for both strains were selected and appended to the peptide selection of SARS-CoV-2. Subsequently, an equimolar dilution series of recombinant nucleoproteins from these three viruses was created in (NH_4_)HCO_3_ as a proof of concept. The particular peptides from both Influenza strains ionized even better than those from SARS-CoV-2, highlighting the potential of SISCAPA-LC-MS for multiplexed testing.

### Conclusion and future perspectives

We have demonstrated that SISCAPA in combination with LC-MS can be used for the high throughput detection of SARS-CoV-2 peptides in patient samples transported in most commonly used media, as well as in saliva and plasma, down to a limit of detection comparable to a viral load of Cq 31 on the E-gene, which is the the most recent estimate for live virus and thus transmittability ^1^. RT-qPCR is perfectly suited for detecting low or high viral load at incredible sensitivities and throughput. Therefore, if the outcome is that patients are quarantined upon any viral detection to avoid further spread of the virus, an accurate quantitative measurement of viral load is redundant. Problematically, based on qPCR results, patients can remain positive long after the infectivity period and in these cases the need for a sensitive antigen test to accurately determine viral load is paramount ^32^. Additionally, quantitative accuracy will become considerably more important when detecting viral peptides in plasma for prognostic diagnosis, disease state, or treatment outcome.

Two specific attributes of the peptide immuno-affinity SISCAPA-LC-MS based workflow provide another application that would greatly improve pandemic monitoring: its versatility in terms of multiplexing and its retained sensitivity after patient pooling. We foresee an assay which targets up to a dozen respiratory pathogens and that allows pooling of up to 32 patients per run. Combining up to 500 assays per day with a sample pooling strategy (minimum 30 samples), 15,000 patients’ samples could theoretically be monitored for a dozen pathogens per instrument per day. This enables longitudinal epidemiological monitoring of the spread of pathogens throughout the population. This early warning system could then alert clinical qPCR labs to change their primer selection to a rising thread. If there are problems with qPCR detection or several pathogens spread through the population at the same time, LC-MS could still be scaled up for their specific detection in single patients. Meanwhile increasing the number of MS instruments in the clinic for detecting a number of other biomarkers would assure that the expertise and capacity is at hand before the next pandemic.

## Data Availability

The mass spectrometry MRM and DIA-MS proteomics data have been deposited to the ProteomeXchange Consortium via the Panorama Public partner repository with the dataset identifier, PXD031401. However, direct access can be obtained by clicking on the following link: https://panoramaweb.org/Cov2MS.url.

## Research Funding

This research was funded by grants from the Research Foundation Flanders (FWO): B.V.P (grant number 11B4518N), R.G. (grant number 1S50918N), L.M. (grant number G042518N), and M.D. (12E9716N); A BOF-COVID-19 grant from the University Ghent Special Research Funding (BOF - 01C01920) and a grant from the European Union’s Horizon 2020 Programme under Grant Agreement 823839 (H2020-INFRAIA-2018-1). Work at SISCAPA Assay Technologies Inc. at the University of Victoria was supported in part by an NSERC (Canada) grant awarded to Dr. Caroline Cameron. A.M. is supported by a PhD student fellowship from the FWO and T.V.R by FWO EOS project VIREOS granted to X.S. The authors thank Dr. Cameron for her advice and contributions to our research. Andrea Bhangu-Uhlmann and Florian C. Sigloch from Polyquant GmbH are acknowledged for providing us with the Cov-MS QconCAT.

