## Supplemental information for "Cov^2^MS: an automated matrix-independent assay for mass spectrometric detection and measurement of SARS-CoV-2 nucleocapsid protein in infectious patients"

#### Supplemental Figures

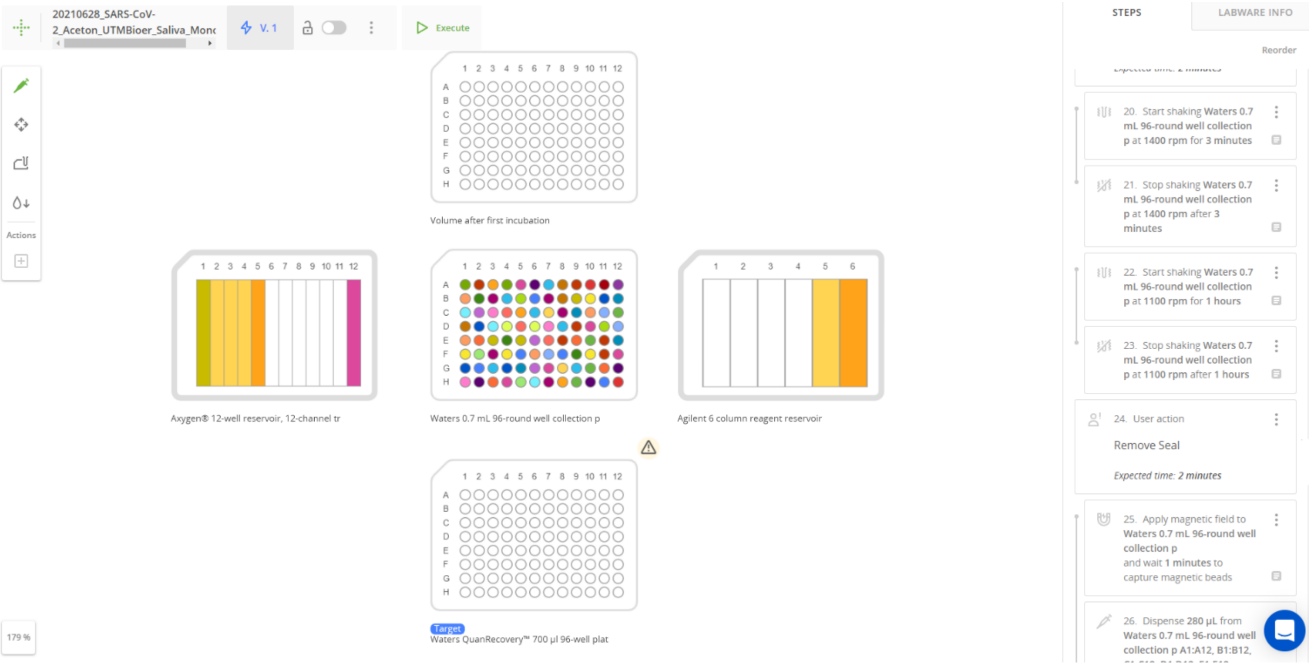


**Supplemental Figure 1.** Screenshot of the protocol automation in the OneLab interface

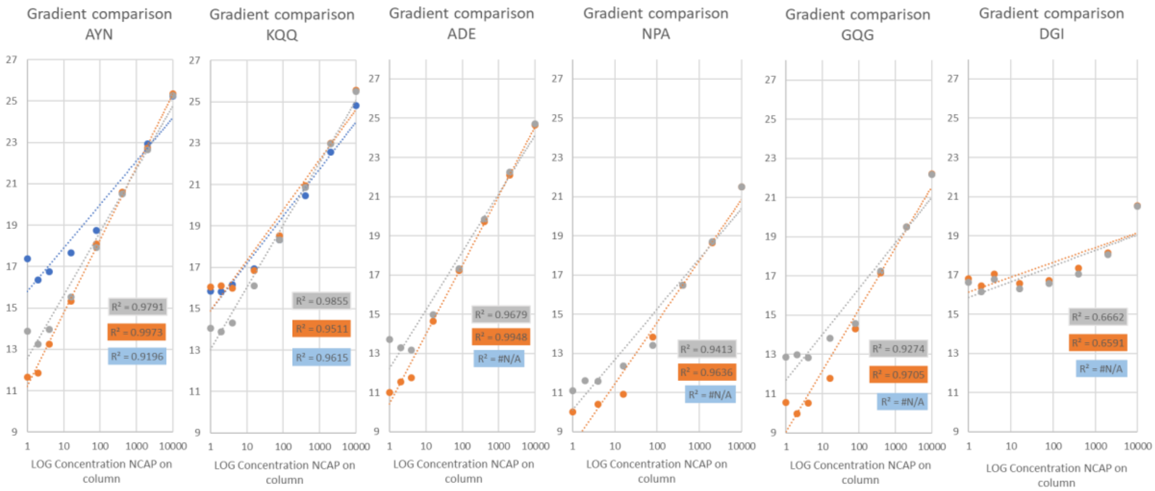


**Supplemental Figure 2.** Response of six SISCAPA peptide targets in a PBS dilution series measured using three different gradients (Blue: 1 min, Orange: 2 min and Grey: 8 min). Here, the x- and y-axis represent the LogConcentration of Nucleoprotein and the corresponding summed peptide LogInt of the MS signal, respectively.

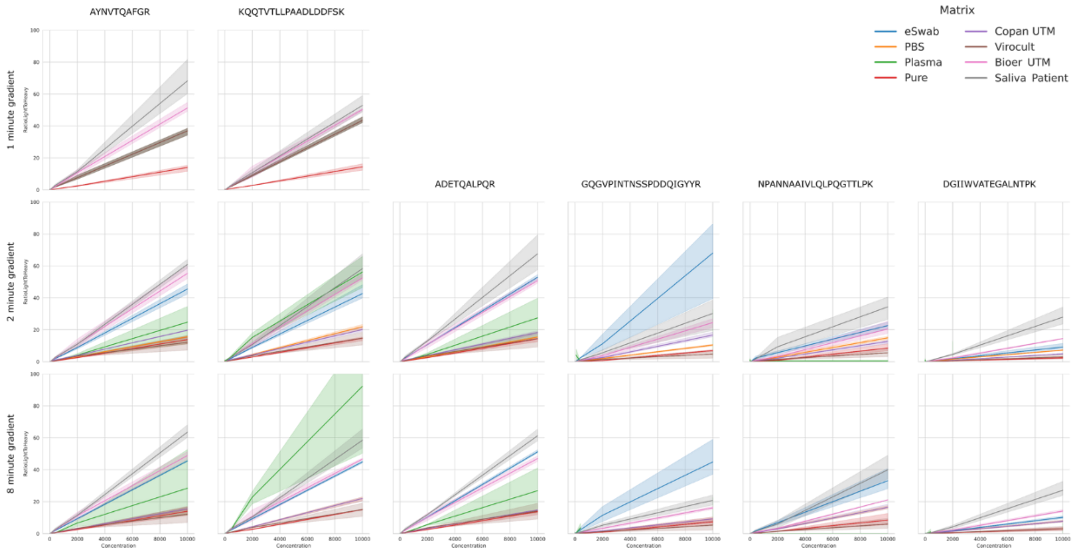


**Supplementary Figure 3.** Light/heavy ratio of six SISCAPA peptides in eight different matrices (i.e. eSwab, PBS, Copan UTM, Virocult, Bioer UTM, Plasma, 100 mM $(NH_4)HCO_3$ and Saliva Patient) using three different gradients.

### Supplementary Figure 4: Gene and peptide selection

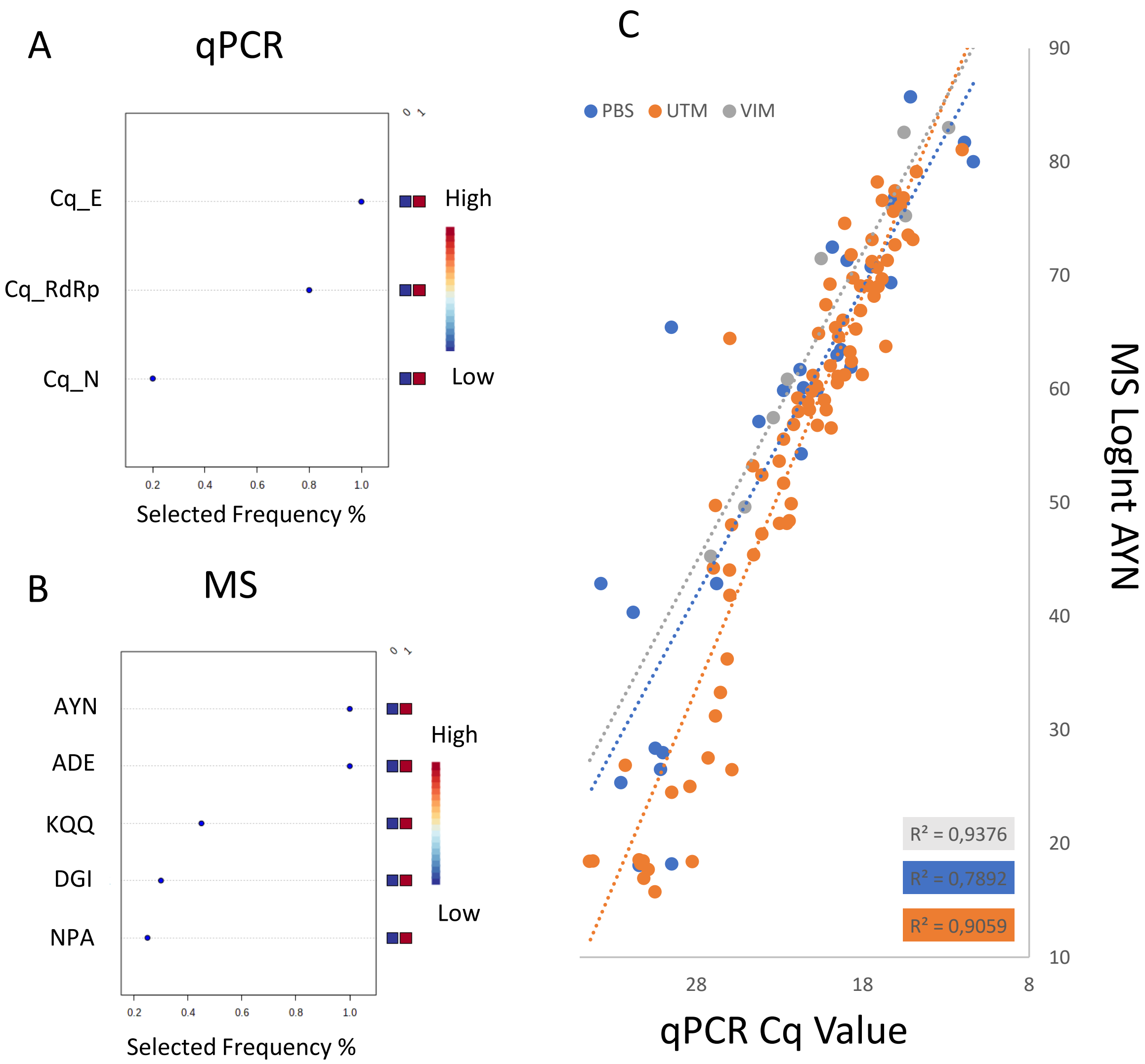


**Supplemental Figure 4.** Contribution of the different **A**) genes (qPCR) and **B**) peptides (MS) to diagnosis as expressed in Selected Frequency % (SF%). **C**) Linear correlation between summed MRM LogInt AYN and Cq for the different media (PBS, UTM and VIM) in the sample batch separately.

**Supplemental Table 1.** MRM parameters for the six SISCAPA target peptides. Transitions highlighted in orange are used as stable isotope labelled standard.

| Peptide | MRM | Cone Voltage (V) | Collision Energy (V) | Retention time (1min) | Retention time (2min) | Retention time (8min) | Scan window (1min) | Scan window (2min) | Scan window (8min) |
|---|---|---|---|---|---|---|---|---|---|
| ADETQALPQR | 564.8 > 712.4 (y6) | 35 | 24 | | 0.48 | 1.13 | | 0.1-0.7 | 0.5-1.6 |
| | 564.8 > 584.4 (y5) | 35 | 20 | | 0.48 | 1.13 | | 0.1-0.7 | 0.5-1.6 |
| | 564.8 > 400.2 (y3) | 35 | 17 | | 0.48 | 1.13 | | 0.1-0.7 | 0.5-1.6 |
| | **572.3 > 407.2 (y3)** | 35 | 17 | | 0.48 | 1.13 | | 0.1-0.7 | 0.5-1.6 |
| AYNVTQAFGR | 563.8 > 892.5 (y8) | 35 | 17 | 0.4 | 0.72 | 2.43 | 0.15-0.8 | 0.5-0.9 | 1.9-2.9 |
| | 563.8 > 778.4 (y7) | 35 | 17 | 0.4 | 0.72 | 2.43 | 0.15-0.8 | 0.5-0.9 | 1.9-2.9 |
| | 563.8 > 679.4 (y6) | 35 | 20 | 0.4 | 0.72 | 2.43 | 0.15-0.8 | 0.5-0.9 | 1.9-2.9 |
| | 563.8 > 349.2 (b3) | 35 | 17 | 0.4 | 0.72 | 2.43 | 0.15-0.8 | 0.5-0.9 | 1.9-2.9 |
| | **571.3 > 689.3 (y6)** | 35 | 20 | 0.4 | 0.72 | 2.43 | 0.15-0.8 | 0.5-0.9 | 1.9-2.9 |
| GQGVPINTNSSPDDQIGYYR | 727.7 > 1126.5 (y9) | 35 | 23 | | 0.81 | 2.62 | | 0.6-1 | 2.1-3.1 |
| | 727.7 > 558.3 (y4) | 35 | 23 | | 0.81 | 2.62 | | 0.6-1 | 2.1-3.1 |
| | 727.7 > 563.8 (y9++) | 35 | 23 | | 0.81 | 2.62 | | 0.6-1 | 2.1-3.1 |
| | 727.7 > 342.2 (b4) | 35 | 23 | | 0.81 | 2.62 | | 0.6-1 | 2.1-3.1 |
| | **736.7 > 570.2 (y9++)** | 35 | 23 | | 0.81 | 2.62 | | 0.6-1 | 2.1-3.1 |
| KQQTVTLLPAADLDDFSK | 664.0 > 1078.5 (y10) | 35 | 18 | 0.44 | 1.18 | 3.8 | 0.15-0.8 | 1-1.4 | 3.3-4.3 |
| | 664.0 > 539.8 (y10++) | 35 | 14 | 0.44 | 1.18 | 3.8 | 0.15-0.8 | 1-1.4 | 3.3-4.3 |
| | 664.0 > 799.5 (b7) | 35 | 22 | 0.44 | 1.18 | 3.8 | 0.15-0.8 | 1-1.4 | 3.3-4.3 |
| | **671.3 > 545.2 (y10++)** | 35 | 14 | 0.44 | 1.18 | 3.8 | 0.15-0.8 | 1-1.4 | 3.3-4.3 |
| NPANNAAIVLQLPQGTTLPK | 687.4 > 841.5 (y8) | 35 | 17 | | 1.19 | 3.9 | | 1-1.4 | 3.4-4.4 |
| | 687.4 > 766.4 (b8) | 35 | 23 | | 1.19 | 3.9 | | 1-1.4 | 3.4-4.4 |
| | 687.4 > 433.2 (b9) | 35 | 20 | | 1.19 | 3.9 | | 1-1.4 | 3.4-4.4 |
| | **696.0 > 851.4 (y8)** | 35 | 17 | | 1.19 | 3.9 | | 1-1.4 | 3.4-4.4 |
| DGIIWVATEGALNTPK | 842.9 > 286.1 (b3) | 35 | 30 | | 1.24 | 4.15 | | 1-1.7 | 3.7-5 |
| | 562.3 > 700.4 (y7) | 35 | 10 | | 1.24 | 4.15 | | 1-1.7 | 3.7-5 |
| | 562.3 > 700.4 (y13++) | 35 | 18 | | 1.24 | 4.15 | | 1-1.7 | 3.7-5 |
| | **852.4 > 289.1 (b3)** | 35 | 30 | | 1.24 | 4.15 | | 1-1.7 | 3.7-5 |
